# A patient satisfaction survey and educational package to improve the care of people hospitalised with COVID-19: an observational study, Liverpool, UK

**DOI:** 10.1101/2021.03.23.21253630

**Authors:** Muhammad Shamsher Ahmed, Scott Rory Hicks, Rebecca Watson, Rajia Akter Ahmed, Lewis Jones, Marcella Vaselli, Meng-San Wu, Fatima Hayat, Libuse Ratcliffe, Mark McKenna, Paul Hine, Sylviane Defres, Tom Wingfield

## Abstract

The experiences of people hospitalised with COVID-19 are under-researched. We designed a COVID-19 patient satisfaction survey and collected responses (n=94) during Liverpool’s first wave (April-June 2020). Although care was generally rated highly, including among people of BAME background, sleep-quality and communication about medications and discharge-planning were identified as areas for improvement. In response, we implemented an education and training package for healthcare professionals working on COVID-19 wards. During Liverpool’s second/third COVID-19 wave, survey responses (n=101) suggested improvement in patient satisfaction across all care domains except discharge-planning and sleep-quality. These UK-first findings are informing local strategies to improve COVID-19 care.

## Introduction

Severe acute respiratory syndrome coronavirus 2 (SARS-CoV-2) has infected over 100 million people worldwide and caused over 2.6 million deaths [1]. In the UK, approximately 11% of individuals affected by coronavirus disease 2019 (COVID-19) have required admission to hospital for treatment [2].

Despite a plethora of research related to COVID-19 vaccines, diagnostics, and biomedical treatments, evidence concerning the perspectives of people with COVID-19, especially from vulnerable and Black Asian Minority Ethnic (BAME) groups, is negligible [3]. To address this, we designed and implemented a satisfaction survey of people with COVID-19 admitted during the first wave of Covid-19 infections (March-June 2020) to our large university hospital in Liverpool, UK. The survey findings (published elsewhere [4]) showed that nursing and medical care was rated highly and most (96%) respondents reported that they would recommend our hospital to friends or family. However, the survey also highlighted potential areas for improvement including communication about medications and their side effects and informing patients about discharge plans [4]. To address these shortcomings and improve holistic COVID-19 hospital care, we subsequently implemented a package of complementary interventions on COVID-19 wards in our centre.

Here we report the satisfaction survey findings from the second/third COVID-19 waves in Liverpool (September 2020 - February 2021) and compare them to those of the first wave.

## Methods

This was an unpowered before-and-after observational quality improvement project (QIP) registered with the local Clinical Effectiveness Department. The COVID-19 patient satisfaction survey was developed in collaboration with our centre’s ‘Patient Experience’ team to ensure it was patient-friendly and suitable for people with learning and reading difficulties.

The survey was adapted from existing patient satisfaction surveys and integrated with our centre’s “Friends and Family Test” questions. An open-access version of the survey itself and accompanying standard operating procedure can be found in [4].

The survey was implemented during the first COVID-19 wave (15^th^ March 2020 to 15^th^ June 2020) and second/third waves (21^st^ September 2020 to 6^th^ February 2021). Between these time periods, the first-wave survey findings were presented at Tropical Infectious Disease Unit QIP meetings and centre-wide Patient Experience meetings. A package of complementary interventions was designed and implemented in August-September 2021. The package consisted of: concise feedback and training sessions for healthcare workers on holistic care on the COVID-19 wards; updated COVID-19 patient information leaflets for admission and discharge; and a ‘COVID-19 Practice Pointers Poster’ (see [4]), which was also placed in visible, shared ward areas.

Descriptive analysis summarised overall responses, compared second/third vs first wave responses, and further compared responses by BAME, age, and gender.

## Results

Surveys from 195 respondents, 94 (48%) from the first wave and 101 (52%) from the second/third wave, were collated (Table 1). Compared to the first wave, in the second/third wave median age was higher (64 vs 59 years) and there were more respondents who were female (58% vs 48%), obese (48% vs 38%), active/ex-smokers (49% vs 33%), and had at least one chronic comorbidity (70% vs 67%). The proportion of BAME respondents was the same across both cohorts (10%).

**Table 1:** Sociodemographic and clinical characteristics of survey respondents (n=195)

| Variables | Respondents (%) |  |  |
| --- | --- | --- | --- |
|  | First wave<br>(n=94) | Second/Third<br>wave (n=101) | All<br>(n=195) |
| **Male** | 49 (52) | 42 (42) | 91 (47) |
| **Age, median (IQR)** | 59 (46-72) | 64 (54-73) | 62 (50-73) |
| **BMI, median (IQR)**<br>(n=84/94, 86/101, 170/195)* | 28 (25-32) | 29 (24-34) | 29 (25-33) |
| **Obesity (BMI ≥ 30 kg/m<sup>2</sup>, n=84/94, 86/101, 170/195)\*** | 32 (38) | 41 (48) | 73 (43) |
| **Black, Asian and Minority Ethnicity** | 10 (11) | 10 (10) | 20 (10) |
| **Smoker / ex-smoker** | 31 (33) | 49 (49) | 80 (41) |
| *Health characteristics* |  |  |  |
| **Chronic lung diseases\*\*** | 36 (38) | 12 (12) | 48 (25) |
| **Hypertension** | 29 (31) | 40 (40) | 69 (35) |
| **Chronic cardiovascular diseases\*\*\*** | 15 (16) | 22 (22) | 37 (19) |
| **Diabetes** | 17 (18) | 32 (32) | 49 (25) |
| **Chronic Kidney Disease** | 13 (14) | 16 (16) | 29 (15) |
| **Non-HIV immunosuppression** | 9 (10) | 7 (7) | 16 (8) |
| **HIV positive** | 2 (2) | 1 (1) | 3 (2) |
| **History of DVT / PE** | 1 (1) | 11 (11) | 12 (6) |
| *Number of Comorbidities* |  |  |  |
| **0** | 31 (33) | 31 (31) | 62 (32) |
| **1** | 34 (36) | 20 (20) | 54 (28) |
| **2** | 14 (15) | 34 (34) | 48 (25) |
| **3 or more** | 15 (16) | 16 (16) | 31 (16) |

*Legend: This table shows the number of respondents (%) in the first (n=94), second/third wave (n=101), and overall (n=195). \*BMI was only available or calculable for 170/195 respondents because height and/or weight was not documented for 10 respondents in the first wave (n=84/94) and 15 respondents in the second/third wave (n=86/101). \*\*Chronic lung diseases include asthma, COPD, bronchiectasis, interstitial lung diseases, lung cancer. \*\*\*Chronic cardiovascular diseases include ischaemic heart disease, heart failure, cerebrovascular disease, and peripheral vascular disease. Abbreviations: IQR interquartile range, BMI body mass index, HIV human immunodeficiency virus, DVT deep vein thrombosis, PE pulmonary embolus*

Patient satisfaction was high with overall care rated 4.8/5 on average. Nearly all (95%) respondents reported that they would recommend our hospital to friends and/or family (Figure 1).

**Figure 1:**
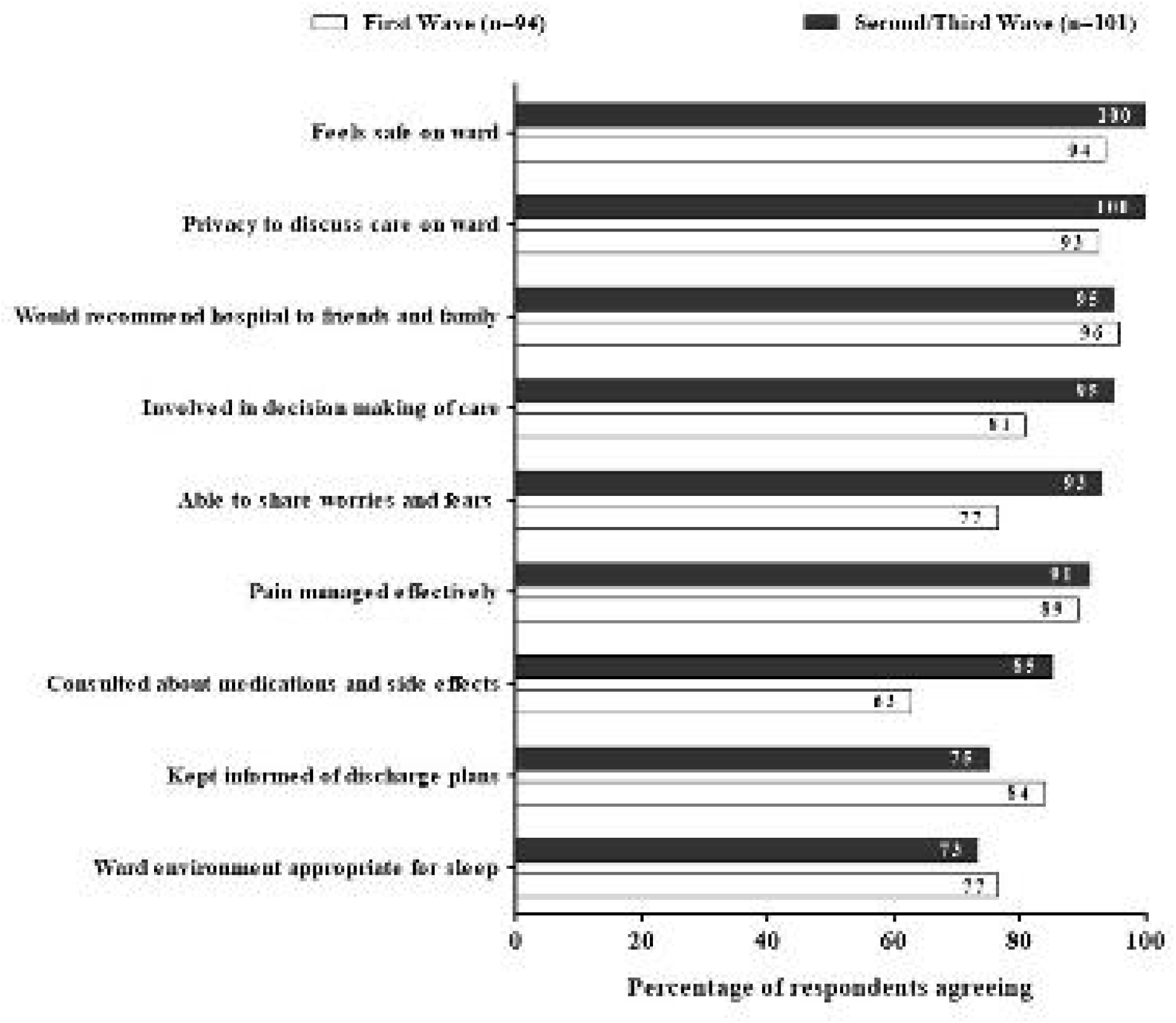
Patient satisfaction survey responses during first and second-third wave of COVID-19 in Liverpool.

Compared to first wave respondents, second/third wave patient satisfaction increased across multiple domains of care but, most notably, being involved in care decisions (81% to 95%), able to share worries and fears (77% to 93%), and communication about medications and side effects (63% to 85%, Figure 1). Satisfaction decreased with relation to being kept informed of discharge plans (84% to 75%) and sleep environment (77% to 73%, Figure 1).

Reported patient satisfaction was higher amongst BAME than non-BAME respondents across all domains except discussion about medications and side-effects (70% vs 75%, Figure 2).Responses were similar by gender and age.

**Figure 2:**
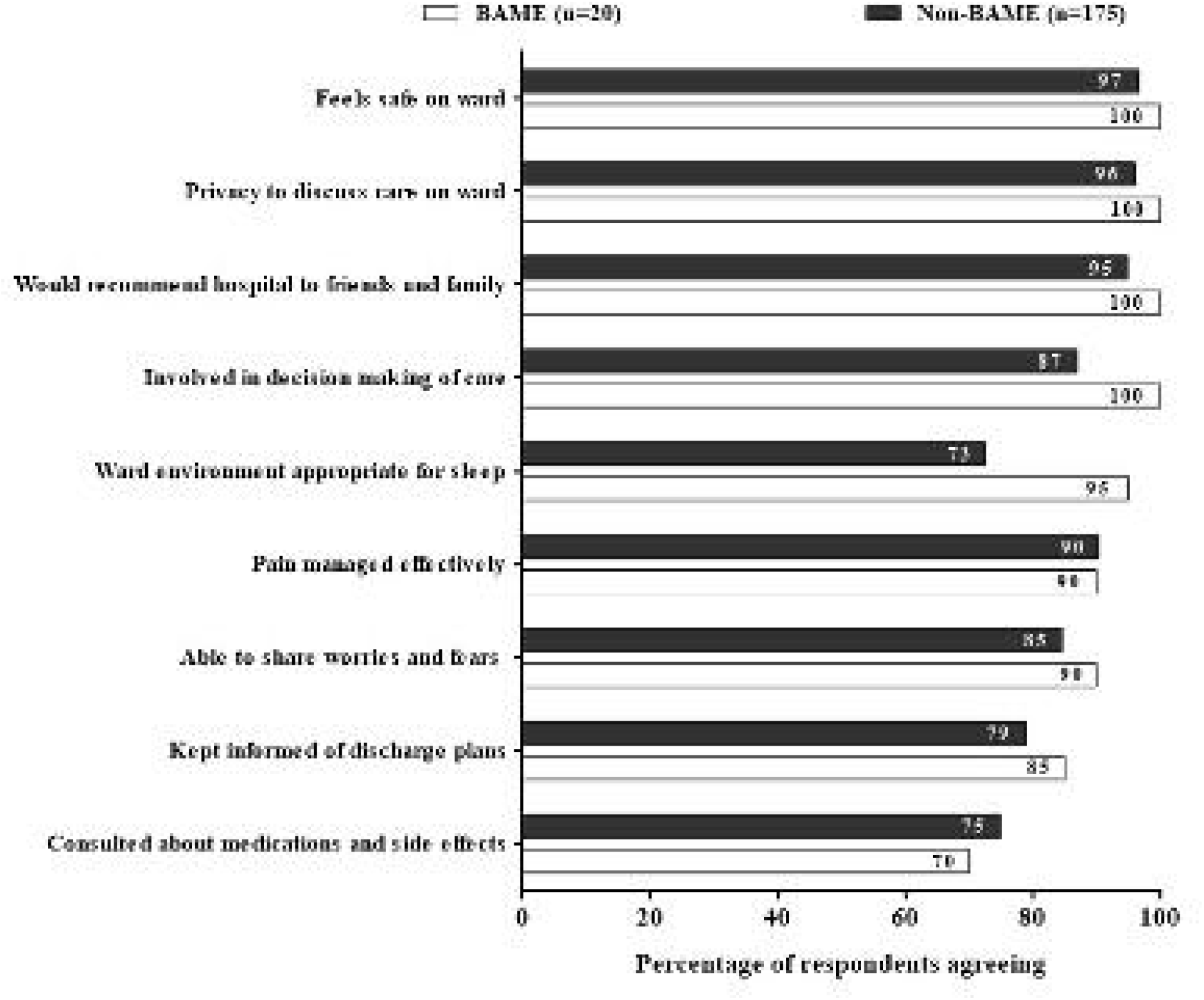
Comparison of patient satisfaction between the BAME and non-BAME respondents.

## Discussion

In the UK to date, nearly half a million people have been hospitalised with COVID-19 [2]. In addition to physical symptoms of COVID-19, those hospitalised can experience negative psychosocial consequences: isolation, including related to infection prevention and control policies; lack of contact with family and friends unable to visit hospital; and uncertainty related to their prognosis [5]. This may be compounded by constrained communication, trust and rapport with healthcare professionals, particularly for people with hearing impairment due to wearing masks and personal protective equipment [3][4][6].

Despite these challenges, our COVID-19 patient satisfaction survey showed that the quality of care at our centre was rated highly throughout the COVID-19 pandemic even during the second/third wave when our local health system was under significant operating pressures. Unfortunately, because this is the first peer-reviewed COVID-19 patient satisfaction survey in the UK, there is currently no comparable regional or national data. However, our findings were consistent with a COVID-19 patient survey from Saudi Arabia [7].

Our finding of higher satisfaction amongst BAME respondents across nearly all care domains was encouraging. Compared to non-BAME, people of BAME background have higher rates of severe COVID-19 disease and death [8] and restricted healthcare access [9]. This serves as a reminder of the importance of addressing the widening health inequalities related to socioeconomic status and ethnicity in the UK [8].

Despite our package of interventions improving some aspects of care [4], sleep quality was rated low across both waves and lower in the second/third wave. These findings align with pre-COVID surveys and are a persistent issue in hospital care [10]. There is ongoing work within our centre to address this.

This was a single-centre, opportunistic, non-randomised survey of a small sample of clinically stable patients, which limits generalisability. Despite this, this study was a UK first and its findings important when considering person-centred COVID-19 care strategies.

## Conclusions

In a cohort of people hospitalised with COVID-19 in Liverpool, hospital care was rated highly including by those of BAME background. Implementation of an education and training package between COVID-19 waves was associated with improved patient feedback, particularly regarding involvement in and communication about care. The survey and package are being expanded locally to further improve care of people with COVID-19 and other conditions.

## Data Availability

Data will be available to review if needed

## Statements

### Contributorship Statement

TW conceived, designed, and supervised the QI project. MSA and TW analysed the data and wrote the manuscript. MSA, SRH, RW, RAA, LJ and MV collected the second/third-wave patient satisfaction data. MW and FH collected the first-wave patient data. MM supported the design of the satisfaction survey. LR, PH and SD supervised the intervention and staff education. All authors reviewed the manuscript and approved the final draft.

